# AI-Assisted Writing Across Medical Journals: Heterogeneity in Prevalence and Trends

**DOI:** 10.64898/2026.10.24.26366055

**Authors:** Michael D Flanders, Michelle Caunca, Ateev Mehrotra, Sarah F Ackley

**Affiliations:** Department of Epidemiology, Brown University, 121 S. Main St., Providence, RI 02903; Department of Neurology, University of California, San Francisco, San Francisco, CA 94118; Department of Health Services, Policy and Practice, Brown University, Providence, RI 02903

## Abstract

**Background:** Estimates of generative artificial intelligence (AI) use in the medical literature vary widely, potentially reflecting differences across journals. We estimated changes in the prevalence of AI-generated writing over time using abstracts from 12 general medical journals.

**Methods:** We randomly sampled up to 200 abstracts per journal-year from publications dated January 1, 2020, through August 8, 2026. Abstracts were scored using GPTZero, with journal-specific thresholds calibrated to fix a 1% false-positive rate (FPR) in the pre-AI baseline period (2020-2021). We estimated annual detector-positive proportions and calculated a publication-volume-weighted estimate across sampled journals. Associations with journal impact factor, publication model, and publication volume were examined descriptively.

**Results:** Among 15,268 abstracts, estimated prevalence of AI-generated text increased after 2022, with substantial variation in timing and magnitude across journals. In 2026 through August 8, prevalence estimates ranged from 5.7% (95% CI, 2.1%-10.0%) in the New England Journal of Medicine to 83.5% (95% CI, 77.1%-87.5%) in Frontiers in Medicine. The publication-volume-weighted estimate across sampled journals was 66.7% (95% CI, 64.0%-69.2%), and trends were insensitive to specification of false-positive rate in baseline data. The proportion of abstracts showing signs of AI-generated text was lower in journals with higher impact factors and higher in fully open-access journals.

**Conclusions:** Estimated prevalence of AI-generated text increased substantially across all sampled medical journals, with marked between-journal differences associated with journal impact factor and publication model. These findings support development of standardized, transparent AI reporting in medical publishing.

## Introduction

Generative artificial intelligence (AI) tools are increasingly common in medical science, but estimates of their use in medical publishing vary widely. Recent work in aggregated corpora has found that 89% of PubMed Central articles showed signs of AI-generated writing in late 2025.^1,2^ Yet studies at leading medical publishers find only about 6% of authors disclosed AI use in a similar period.^3,4^ It is tempting to attribute this discrepancy to uniform underreporting by authors, but it could equally reflect genuine heterogeneity in AI use across journals or publishers. We therefore estimate longitudinal changes in prevalence of AI-assisted writing across 12 general medical journals, using article abstracts to include publications beyond the open-access record.

## Methods

We retrieved abstracts published from January 1, 2020, through August 8, 2026 from 12 medical journals using the NCBI Entrez E-utilities application programming interface (API). From each journal-year stratum, 200 abstracts were selected by random sampling; titles, author information, and other metadata were excluded from the text evaluated. (Journal selection, abstract eligibility criteria, and evaluable abstract counts in Supplemental Methods.)

We scored abstract texts for likely human authorship with the GPTZero API^5^ (model 4.9b, 2026-08-09-base), assigning each abstract a continuous human authorship score. All detectors produce some noise, while style and formatting differ across journals. To address this, for each journal we empirically selected a threshold value that classified 1% of abstracts published in 2020-2021 as detector-threshold-positive (setting a 1% false positive rate in the baseline period preceding ready availability of AI writing tools). Abstracts scoring below the journal-specific threshold were classified positive for evidence of AI writing.

Using these fixed journal-specific thresholds, we estimated the prevalence of AI-text-positive abstracts with 95% CIs for each journal-year by bootstrap resampling with 1000 replicates. We assessed sensitivity of these prevalence estimates to choice of threshold values by repeating the analyses with alternative reasonable baseline period false-positive rates of 2, 3 and 4%. We also examined, at the journal level, associations between the 2026 estimated prevalence of AI text and journal impact factor (Clarivate 2024 Journal Impact Factors), publication model (subscription/hybrid versus fully open access), and 2025 full-year PubMed publication volume. R version 4.5.1 was used for all analyses.

## Results

The analysis included 15268 abstracts from the 12 journals. Across journals, estimated prevalence of abstracts containing AI-generated text was stably near zero before widespread AI availability, then increased after 2022 with varying latency (**Figure 1**). In 2026 through August 8, estimated prevalence ranged from 5.7% (95% CI, 2.1%-10.0%; NEJM) to 83.5% (95% CI, 77.1%-87.5%; Frontiers in Medicine). The publication-volume-weighted prevalence estimate across the 12 journals was 66.7% (95% CI, 64.0% - 69.2%). Estimates were insensitive to baseline threshold specification: across FPRs of 1%-4%, volume-weighted 2026 prevalence ranged from 66.7% to 73.9%. Estimated prevalence was lower at journals with higher impact factors, and higher among fully open-access journals **(Figure 2)**.

**Figure 1.**
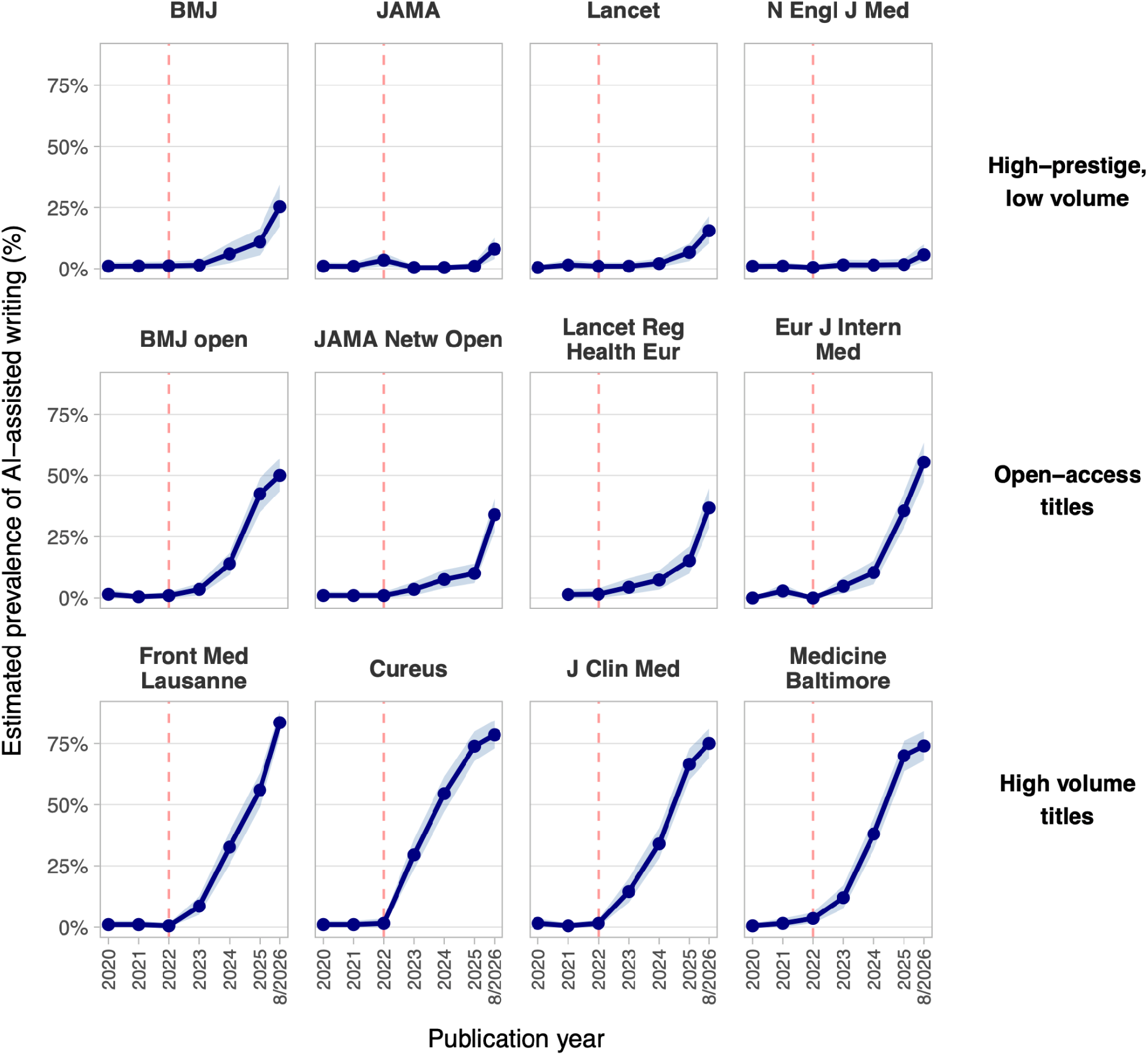
Estimated Prevalence of AI-Generated Text in Medical Journal Abstracts by Year. Points show estimated prevalence of abstracts classified as positive for AI text in a given journal-year; shaded areas represent 95% bootstrap CIs. Dashed red lines denote the public release of the first widespread AI tool, ChatGPT, in November 2022. 2026 is a partial year, representing articles published through August 8 of that year. Abbreviations: BMJ, *British Medical Journal*; JAMA, *Journal of the American Medical Association*; Lancet, *The Lancet*; N Engl J Med, *New England Journal of Medicine*; BMJ open, *BMJ Open*; JAMA Netw Open, *JAMA Network Open*; Lancet Reg Health Eur, *Lancet Regional Health – Europe*; Eur J Intern Med, *European Journal of Internal Medicine*; Front Med Lausanne, *Frontiers in Medicine*; Cureus, *Cureus*; J Clin Med, *Journal of Clinical Medicine*; Medicine Baltimore, *Medicine*.

**Figure 2.**
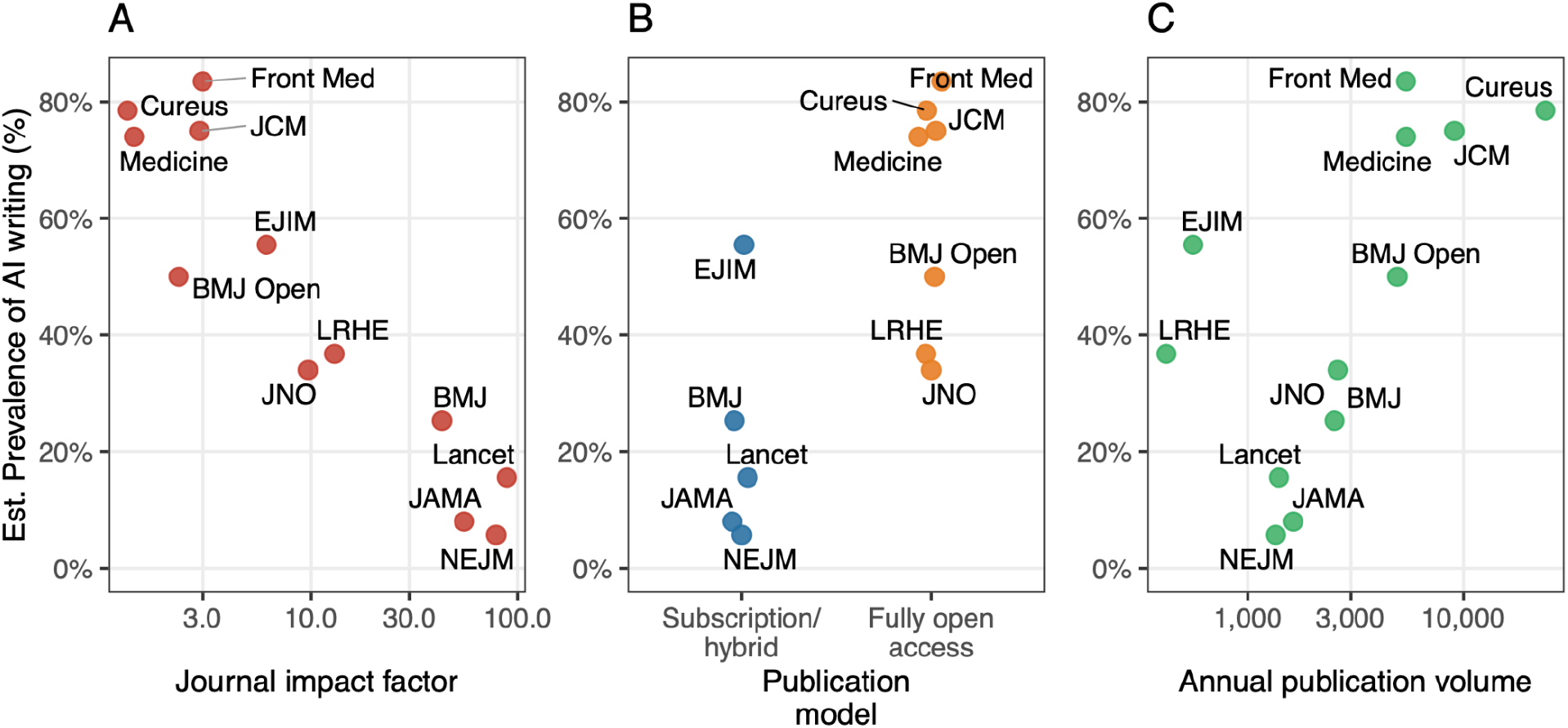
Journal Characteristics and Estimate Prevalence of AI-Generated Text in 2026. Estimated prevalence of AI-generated text January 1 through August 8, 2026, displayed by 2024 journal impact factor (A), publication model (B), and number of PubMed-indexed articles published in all of 2025 (C). Impact factor and annual publication volume are displayed on logarithmic scales. Journal-level comparisons are descriptive. Abbreviations: BMJ, *British Medical Journal*; JAMA, *Journal of the American Medical Association*; Lancet, *The Lancet*; NEJM, *New England Journal of Medicine*; BMJ Open, *BMJ Open*; JNO, *JAMA Network Open*; LRHE, *Lancet Regional Health – Europe*; EJIM, *European Journal of Internal Medicine*; Front Med, *Frontiers in Medicine*; Cureus, *Cureus*; JCM, *Journal of Clinical Medicine*; Medicine, *Medicine*

## Discussion

In this longitudinal analysis of 12 journals, two-thirds of medical journal abstracts contained signs of AI writing in 2026. This volume-weighted estimate agrees in magnitude with recent findings.^2^ However, we find striking differences across sampled medical journals both in the current prevalence of AI-generated writing and in the apparent rate of AI adoption. These differences appear related to journal impact factor, publication volume, and publication model.

This study has limitations. GPTZero is a proprietary AI detection tool, and our approach to its use deliberately does not distinguish modest use of AI from wholly AI-generated text.

Journal-specific calibration reduces confounding from stable stylistic differences, but changes in article mix, authorship, or editorial practice over time could contribute to these trends. This analysis does not distinguish signals of AI arising in authors’ manuscript preparation from those that could arise during journals’ editorial processing. The presence of AI-generated text in an abstract does not necessarily reflect use of AI in the corresponding full text, though these appear strongly related in aggregate approaches.^2^ Given the number of journals assessed, comparisons in Figure 2 should be viewed as descriptive. Finally, because AI detection methods lag behind developments in generative AI tools, recent prevalence is likely underestimated.^6^

These findings suggest that AI-generated text is now common in medical publishing, but is very unevenly distributed. This informs interpretation of the contemporary medical literature because, while AI provides clear gains in productivity, recent work suggests it also carries risks to the integrity of the scientific record.^7–10^ Standardized and transparent reporting of AI assistance will be essential to understanding how AI tools are changing the production and interpretation of medical evidence.

## Data Availability

All data produced in the present study are available upon reasonable request to the authors.

## Acknowledgements

The authors thank GPTZero for providing qualified researchers with free credits and API access to detection models via their public-facing Scholarly Access program at https://gptzero.me/.

## Author Contributions

Concept and design: MDF. Acquisition, analysis, or interpretation of data: MDF, SFA. Drafting of the manuscript: MDF, MC, AM. Critical revision of the manuscript for important intellectual content: MC, AM, SFA. Statistical analysis: MDF, SFA. Supervision: SFA.

## Data Access and Responsibility

MDF and SFA had full access to all the data in the study and take responsibility for the integrity of the data and the accuracy of the data analysis.

## Conflict of Interest Disclosures

MDF reported no disclosures. MC reported funding from the National Institute of Neurological Disorders and Stroke (5UE5NS070680-17 and K23NS151370) and the Alzheimer’s Association (P0590624). AM reported serving on paid advisory boards for Black Opal Ventures and Perplexity AI. SFA reported receiving funding to Brown University from the National Institute on Aging (R00AG073454) and from Sanofi for work unrelated to AI in medical research.

## Funding/Support

This study received no specific funding.

## Role of the Funder/Sponsor

N/A

## AI use

ChatGPT 5.6 (OpenAI; used September 2026) was used to assist with condensation of a human-generated manuscript, and Claude Sonnet 4.6 (Anthropic; used September 2026) was used to refactor human-generated and -tested analytic code. All manuscript content was reviewed by the authors, and code was reviewed by MDF and SFA. The authors take responsibility for the integrity of all AI-assisted content.

## Notes

### Competing Interest Statement

MDF: no competing interests. MC: funded by NINDS (5UE5NS070680-17 and K23NS151370) and the Alzheimer's Association (P0590624). AM: on paid advisory boards for Black Opal Ventures and Perplexity AI. SFA: receives funding to Brown University on her behalf from NIH NIA (R00AG073454) and from Sanofi for work unrelated to the use of AI in medical research.

## References

1. Kobak D, González-Márquez R, Horvát EÁ, Lause J. Delving into LLM-assisted writing in biomedical publications through excess vocabulary. Sci Adv. 2025;11(27):eadt3813.

2. Holzwarth L, González-Márquez R, Kobak D. Most biomedical publications show signs of LLM-assisted writing. arXiv [csCL]. Published online August 11, 2026. doi:10.48550/arXiv.2608.10715

3. AlFayyad I, Zeegers MP, Bouter L, Macdonald H, Schroter S. Self-disclosed use of AI in research submissions to BMJ journals. JAMA. 2026;335(8):719–721.

4. Perlis RH, Flanagin A, Kendall-Taylor J, Berkwits M, Bibbins-Domingo K. Author disclosure of use of AI in submissions to 13 JAMA network journals. JAMA. 2026;335(8):717–719.

5. Adam GA, Cui A, Thomas E, et al. GPTZero: Robust detection of LLM-generated texts. arXiv [csLG]. Published online February 13, 2026. doi:10.48550/arXiv.2602.13042

6. Elkhatat AM, Elsaid K, Almeer S. Evaluating the efficacy of AI content detection tools in differentiating between human and AI-generated text. Int J Educ Integr. 2023;19(1):17.

7. Topaz M, Roguin N, Gupta P, Zhang Z, Peltonen LM. Fabricated citations: an audit across 2·5 million biomedical papers. Lancet. 2026;407(10541):1779–1781.

8. Stokel-Walker C. Scientists invented a fake disease. AI told people it was real. Nature. 2026;652(8110):559–561.

9. Omar M, Sorin V, Wieler LH, et al. Mapping the susceptibility of large language models to medical misinformation across clinical notes and social media: a cross-sectional benchmarking analysis. Lancet Digit Health. 2026;8(1):100949.

10. Tordjman M, Yuce M, Ammar A, et al. The rise of deepfake medical imaging: Radiologists’ diagnostic accuracy in detecting ChatGPT-generated radiographs. Radiology. 2026;318(3):e252094.

