## Supplemental Methods for "AI-Assisted Writing Across Medical Journals: Heterogeneity in Prevalence and Trends"

**Journal Selection and Data Collection.** Journals were selected to span publication models, impact factors, and publication volumes and included 4 historically preeminent general medical journals, their open-access counterparts where available, and 4 high-volume medical journals. The sample was not intended to represent all medical literature. Abstracts containing 140 to 600 words were considered evaluable; records outside this range were commonly errata or other nonstandard records. Sampling within each journal-year stratum used a fixed random seed (123). PubMed records were retrieved through the NCBI Entrez E-utilities API using the R package rentrez.

**AI-Authorship Detection.** Abstract text was scored using the GPTZero API (model 4.9b, 2026-08-09-base), which assigns a continuous estimated probability of human authorship. API errors were excluded from analysis. The primary detection measure was the human-authorship score. Only abstract text was submitted to the detector; titles, author information, and other metadata were excluded. Counts of evaluated abstracts per journal-year stratum are shown in Table S1.

**Classification Threshold Derivation**. Because writing style and abstract format differ across journals, journal-specific thresholds were derived from the distribution of human-authorship scores among abstracts published in 2020–2021 (before the public release of ChatGPT in November 2022). For the primary analysis, the threshold for each journal was set at the first percentile of its baseline score distribution, corresponding empirically to a 1% baseline false-positive rate. Abstracts with scores below this threshold were classified as positive for evidence of AI writing. Journal-specific thresholds are shown in Table S2.

**Primary and Sensitivity Analyses**. Using these journal-specific thresholds, we estimated the prevalence of AI-text-positive abstracts with 95% CIs for each journal-year by bootstrap resampling with 1000 replicates. Sensitivity analyses repeated threshold calibration using baseline-period false-positive rates 1%, 2%, 3% and 4%. At the journal level, we also descriptively examined associations between estimated 2026 AI-text prevalence and 2024 journal impact factor (used to permit inclusion of *Cureus*, which was subsequently delisted from Clarivate JCR), publication model (subscription/hybrid vs fully open access), and 2025 full-year PubMed publication volume. Given 12 journals analyzed, these comparisons are descriptive in nature. All analyses and orchestration were conducted using R version 4.5.1.

**Table S1. Number of Evaluable Abstracts Sampled per Journal-Year Stratum.** The target sample was 200 abstracts per journal-year stratum. Lower-volume journals did not always have 200 abstracts meeting eligibility criteria. *Lancet Reg Health Eur* had <10 evaluable abstracts in year 2020.

| | **2020** | **2021** | **2022** | **2023** | **2024** | **2025** | **→ Aug 8 2026** |
|---|---|---|---|---|---|---|---|
| **BMJ** | 187 | 177 | 168 | 138 | 132 | 128 | 87 |
| **JAMA** | 191 | 200 | 200 | 199 | 200 | 193 | 150 |
| **Lancet** | 200 | 193 | 192 | 190 | 189 | 194 | 135 |
| **N Engl J Med** | 200 | 188 | 188 | 189 | 194 | 174 | 140 |
| **BMJ open** | 200 | 200 | 200 | 200 | 200 | 200 | 200 |
| **JAMA Netw Open** | 198 | 199 | 200 | 200 | 200 | 200 | 200 |
| **Lancet Reg Health Eur** | — | 142 | 129 | 138 | 178 | 171 | 136 |
| **Eur J Intern Med** | 177 | 141 | 133 | 168 | 145 | 163 | 137 |
| **Front Med Lausanne** | 200 | 200 | 200 | 199 | 199 | 200 | 200 |
| **Cureus** | 200 | 200 | 200 | 200 | 200 | 199 | 200 |
| **J Clin Med** | 200 | 200 | 200 | 200 | 200 | 200 | 200 |
| **Medicine Baltimore** | 200 | 200 | 200 | 200 | 200 | 200 | 200 |

**Table S2. Journal-Specific Human-Authorship Score Thresholds Corresponding to a 1% Baseline False-Positive Rate.** Thresholds were derived separately for each journal from abstracts published in 2020-2021 to account for baseline differences in writing style and abstract structure.

| | 1% FPR threshold |
|---|---|
| BMJ | 0.981 |
| JAMA | 0.903 |
| Lancet | 0.946 |
| N Engl J Med | 0.991 |
| BMJ open | 1.000 |
| JAMA Netw Open | 0.975 |
| Lancet Reg Health Eur | 0.993 |
| Eur J Intern Med | 0.995 |
| Front Med Lausanne | 0.992 |
| Cureus | 0.823 |
| J Clin Med | 0.959 |
| Medicine Baltimore | 0.994 |
